# Beyond External Load: Integrative Immune Monitoring Reveals Injury-Predictive Signals in the Athlete’s Internal State

**DOI:** 10.64898/2026.06.06.26354898

**Authors:** Umberto Perron, Ana Mendizabal-Sasieta, Marta Grzelak, Marta Soto, Mariana Capelli, Andres Martin-Garcia, Milos Mallol, Gil Rodas, Ricard Pruna, Lucas Gómez-Chereguini, Holger Heyn

## Abstract

Injury risk prediction in elite football relies almost exclusively on external load metrics derived from GPS tracking, overlooking the molecular state of the athlete. We monitored 26 male players from FC Barcelona’s first team across the 2025 calendar year, integrating GPS-derived training load with longitudinal blood-based immune monitoring (systemic inflammation and TCR-derived immune age). Immune age acceleration and inflammation were elevated in the 14 days preceding musculoskeletal injuries. A logistic regression model combining external load, inflammation, immune age acceleration, and career injury history reached an overall AUC of 0.678 and a mean per-player AUC of 0.754 (SD 0.146), improving on a GPS-only baseline of 0.541. Applied to 2026 data, the frozen model ranked players who later sustained non-contact musculoskeletal injuries high in the risk distribution.

Together, our data suggest multimodal immune monitoring in elite football to reveal the athlete’s internal physiological state, which carries injury-relevant information that external load alone does not capture.

## 1. Introduction

Muscle injuries are a primary issue in elite football despite a declining trend in overall injury incidence across UEFA competitions [1]. Hamstring injuries alone now constitute 24% of all injuries in the UEFA Elite Club Injury Study, with a median absence of 13 days and an 18% recurrence rate [2]. The economic and competitive consequences are quantified in Section 3.1.

Current approaches to injury prediction rely predominantly on the acute:chronic workload ratio (ACWR) framework, which quantifies the relationship between recent and background training load from external load metrics, commonly derived from GPS tracking [3]. While ACWR has been widely adopted in professional sport, its validity as a standalone predictor is increasingly questioned due to mathematical coupling artefacts that inflate apparent predictive power [4]. Beyond ACWR, machine-learning-based injury prediction in sport faces methodological challenges: a recent scoping review of 38 ML injury-prediction studies in sport reported AUCs ranging from 0.57 to 0.95, with only 18% applying any form of model explainability and 53% of studies relying solely on preseason screening tests. Here, only 18% of studies analysed external load data from GPS tracking, and broader physiological or molecular monitoring was rarer still [5].

Blood biomarkers are routinely measured in professional football for workload management and health monitoring [6]. Studies combining standard blood panel features (e.g., haematocrit, testosterone, cortisol, creatine kinase) [16] or broader panels of up to 40 biomarkers [17] with GPS-derived workload data have shown modest improvements over load-only baselines. However, high-throughput molecular profiling (e.g. transcriptomics, proteomics) has not yet been integrated with GPS-based injury risk models. The 2024 MoTrPAC Consortium’s multi-omic exercise atlas has revealed extensive immune, metabolic, and mitochondrial pathway regulation in blood in response to physical activity in animal models [7], which suggests that these signals could be exploited for injury prediction if profiled at sufficient temporal resolution. Parallel evidence demonstrates that: 1) acute exercise remodels the PBMC proteome, altering over 6,000 proteins across immune effector pathways [8]; 2) that transcriptomic signatures correlate with GPS-derived external load in competitive handball [9]; and 3) that longitudinal multi-omic profiling during high-altitude mountaineering reveals adaptive and maladaptive immune responses at different exercise intensities [10].

These observations align with a broader concept of *internal load biology* : external load metrics quantify an athlete’s mobility, but they do not describe how the body responds to this stimulus. Targeted metabolomic profiling of professional footballers, for example, has shown that metabolic adaptation to player load differs between sexes and is only partly explained by session-level external load [24], supporting the principle that a meaningful fraction of the training stimulus is encoded in the athlete’s internal molecular state rather than in the workload measurement itself.

Taken together, these findings suggest that systemic inflammation and TCR-derived immune age may provide an independent and biologically informative signal for injury susceptibility, one that reflects the athlete’s internal physiological state rather than the external training stimulus.

Our objective with this study was twofold: 1) to characterise the relationship between systemic inflammation, immune dynamics, training load, and injury risk in elite male footballers; and 2) to develop and evaluate an injury risk prediction model integrating GPS load metrics, inflammation, immune age acceleration, and career injury history.

## 2. Methods

### 2.1 Study design and cohort

This was a retrospective longitudinal cohort study of 26 male professional football players from FC Barcelona’s (FCB) first team, monitored continuously across the 2024–2025 seasons (January 2025 to December 2025). All players had complete data across four modalities: GPS training sessions, blood-based inflammation profiling, TCR-derived immune age (osAge, Omniscope), and injury records. The study was conducted as part of routine sports science monitoring; all data were collected under the club’s existing medical and performance protocols.

**Figure 1.**
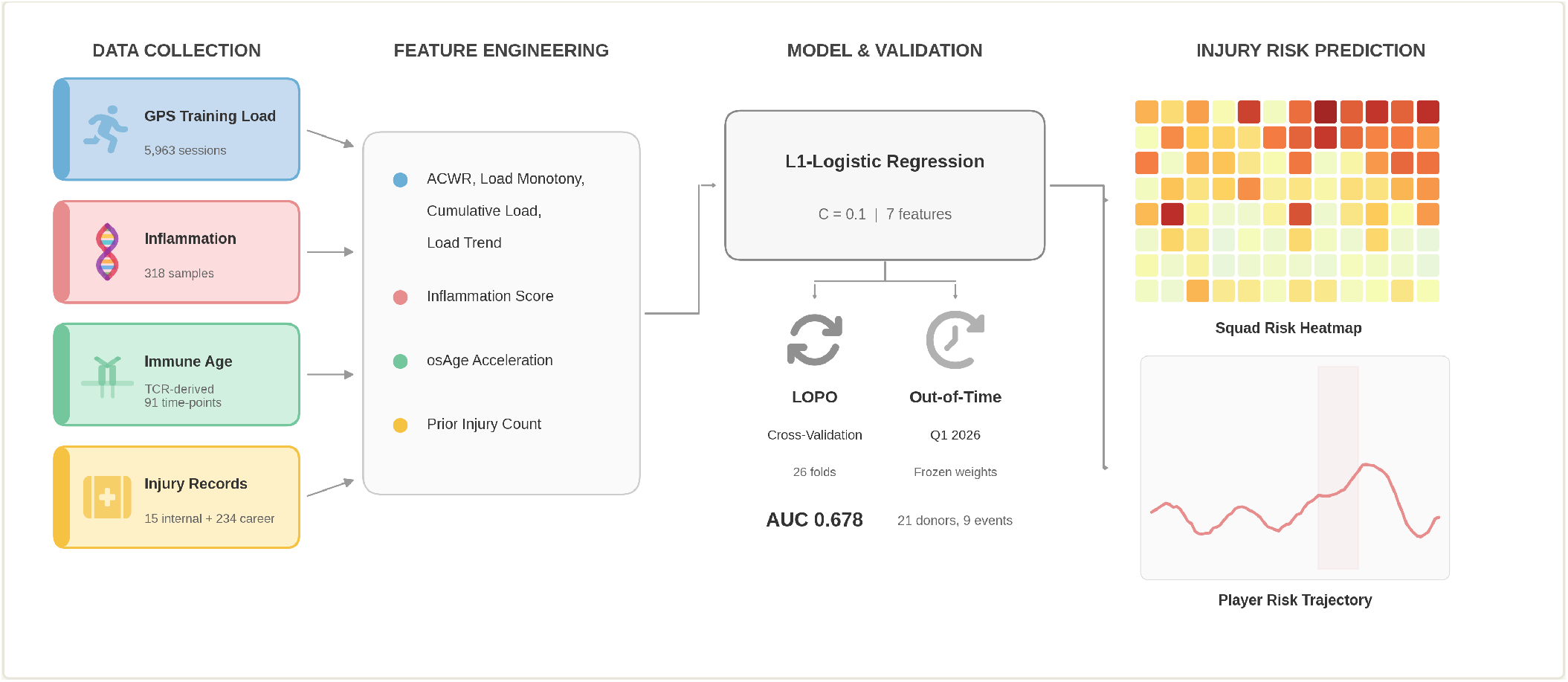
Study design schematic. Longitudinal data collection across four modalities from 26 FCB first-team players (2024–2025): GPS training load (5,963 sessions), inflammation profiling based on peripheral blood (318 samples), TCR-derived immune age predictions (91 assessments), and injury records (15 internal events, 234 career history events). Features are merged by temporal alignment and fed into an L1-regularised logistic regression model with leave-one-player-out cross-validation.

### 2.2 Data sources

#### GPS and wearables

Training session data were collected via GPS-embedded inertial measurement units, yielding 5,963 sessions across the cohort. The primary load metric was Player Load (PL, arbitrary units), a triaxial accelerometry-derived measure of external load. Additional metrics included high-speed running distance (HSR), high metabolic load distance (HMLD), and accelerometer-derived acceleration and deceleration counts.

#### Inflammation

Systemic inflammation was profiled from 318 longitudinal peripheral blood samples using a bulk transcriptomics processing pipeline (alignment, normalisation, and batch correction [11,12,18]). Per-sample inflammation was summarised into a single enrichment score (ESn) using Omniscope’s Inflammation Score, a proprietary analytics service that quantifies coordinated regulation of functionally-related gene sets as a readout of the inflammatory response to stress.

#### Immune age

A TCR-derived (osTCR, Omniscope [29]) immune-age acceleration feature was generated by ImmuneGPT (Omniscope; USPTO Serial No. 99410386), a proprietary model that reads out an immune biological age (osAge) from an individual’s T-cell receptor repertoire. Immune age acceleration was defined as osAge minus the player’s chronological age. ImmuneGPT is used here as a fixed upstream feature generator; its weights are not updated using the injury outcomes, which limits the risk of outcome-driven overfitting in the molecular branch. A total of 91 predictions were available across 25 athletes, with a matching window of 90 days to the nearest GPS session.

#### Injury events

Two sources of injury data were integrated. FCB’s internal injury cards documented 15 musculoskeletal events from 12 athletes during the 2025 study period: 12 muscle injuries of the lower limb (predominantly biceps femoris and soleus), 2 knee injuries, and 1 general injury, of which 12 (80%) were classified as non-contact. Across all sources (UEFA cards plus Transfermarkt career-history events restricted to the same window), the cohort sustained 68 documented injury events in 2025, of which 36 were muscle injuries. The 15 internally documented events thus represent approximately 22% of the 2025 squad’s total injury burden, and the 12 muscle subset represents approximately 33% of the season’s muscle-injury burden. UEFA cards were extracted from injury documents via direct XML parsing, providing precise injury dates, anatomical locations, and contact/non-contact classification. All diagnoses were performed by the same FCB medical staff throughout the study period, with ultrasound and magnetic resonance imaging used where indicated; criteria for diagnosis, prognosis, and return-to-play followed the MLG-R muscle-injury classification developed at FCB [27]. External career histories (234 injuries across 26 players) were manually retrieved from Transfermarkt and curated for the prior injury count feature. Because Transfermarkt is a public, non-medical source with limited temporal precision (estimated ±3–5 days) and coarse injury-type classification, it is used here only as a contextual/exploratory layer to enrich a single career-history feature, not as a primary outcome label. Both sources were harmonised with a 14-day deduplication window, yielding 169 combined events. Internal injuries served as prediction labels; combined data enriched the prior injury count feature only.

### 2.3 Feature engineering

Features were engineered across the four modalities: external training-load metrics derived from GPS (including the acute:chronic workload ratio, load monotony, and cumulative and trend load [3]), prior musculoskeletal injury count from career history, a TCR-derived immune-age acceleration feature, and a blood inflammation feature. GPS features were computed per training session; molecular features were matched to GPS sessions using nearest-date temporal alignment within their respective windows, and sessions without a match were excluded.

### 2.4 Model specification

An L1-regularised (Lasso) logistic regression model was trained with regularisation strength C = 0.1 using scikit-learn v1.7.0 (LogisticRegression, saga solver). The binary outcome was defined as any internally annotated musculoskeletal injury occurring within the subsequent 14 days. L1 regularisation was chosen to provide automatic feature selection and coefficient shrinkage appropriate for the small event count, while maintaining coefficient interpretability. No feature selection was performed outside the regularisation.

Cross-validation followed a leave-one-player-out (LOPO) design: in each of 26 folds, the model was trained on data from 25 players and evaluated on the held-out player. This prevents within-player data leakage and provides a realistic estimate of generalisation to new athletes. Features were standardised within each training fold.

### 2.5 Statistical analysis

Pre-injury inflammation was compared between positive (injury within 14 days) and negative observations using Welch’s t-test, which accounts for unequal group sizes (n = 116 vs n = 3,426). Effect size was quantified with Cohen’s d. Model discrimination was assessed by overall AUC-ROC and per-player AUC (computed for the 12 players with at least one positive observation). Calibration was assessed by Brier score. Sensitivity, specificity, positive predictive value (PPV), and negative predictive value (NPV) were reported at the Youden-optimal operating threshold.

### 2.6 Economic impact estimation

The economic burden of non-contact muscle injuries was estimated in three steps. First, the expected annual injury days per squad were derived from published epidemiological data [2,19]: approximately 15 muscle injuries per 25-player squad per season, of which 92–96% are non-contact depending on muscle group, with a median absence of 13 days (IQR 7–22). We used the upper bound (96%, corresponding to hamstring and quadriceps injuries) and the median rather than the mean absence (18 days for structural injuries) because the absence distribution is right-skewed; using the mean would increase the estimate by approximately 40%.

Daily player compensation was then estimated from Transfermarkt squad market values (2024–25 season), which have been validated as proxies of player salary (log-log r ≥ 0.70 against actual wages in MLS data where salaries are public [23]). The relationship between market value and wages is non-linear and tier-dependent; we used a simplified conversion at 50% of squad market value as the estimated annual wage bill, acknowledging that this ratio overstates compensation for the highest-valued players and understates it for lower tiers [23]. Daily cost was computed as estimated annual wage bill / 25 players / 365 days. Three club tiers were defined by squad market value (via Transfermarkt, accessed March 2025): elite (∼EUR 470M; e.g., FC Barcelona, Real Madrid, Manchester City), contender (∼EUR 274M; e.g., Liverpool, Atletico Madrid, Dortmund), and standard (∼EUR 116M; e.g., Villarreal, Freiburg, Fulham). This approach assumes uniform daily cost across the squad; in practice, injuries disproportionately affect high-load starters whose per-day cost exceeds the squad average [20], so actual costs for elite clubs are likely higher than estimated.

Third, illustrative savings scenarios were computed by multiplying hypothetical injury reduction percentages (5%, 10%, 20%) by the estimated annual injury-day cost per tier.

### 2.7 Ethics and consent

The study was approved by the ethics committee of the Barça Innovation Hub (FC Barcelona). All players signed informed consent in pre-season authorising the use of biological, laboratory, and training data for research purposes under the club’s existing medical and performance protocols. No additional procedures beyond routine sports-science monitoring were performed for this study.

## 3. Results

### 3.1 Injury burden and economic context

Muscle injuries constitute the largest single injury category in elite football, with approximately 15 muscle injuries per 25-player squad per season (92–96% non-contact) and a median lay-off of 13 days (IQR 7–22) [19]. Hamstring injuries account for 24% of all injuries and have doubled in incidence over 21 seasons of the UEFA Elite Club Injury Study, with an 18% recurrence rate [2]. Using the approach described in Section 2.6, we estimate 187 non-contact muscle injury days per squad per season.

**Table 1a.** Estimated annual salary cost of non-contact muscle injury days by club tier (see Section 2.6 for derivation).

| CLUB TIER | EXAMPLE CLUBS | EST. DAILY COST<br>PER PLAYER (EUR) | INJURY DAYS/<br>SEASON | ANNUAL COST<br>(EUR) |
| --- | --- | --- | --- | --- |
| Elite | FC Barcelona, Real Madrid, Man City | ~25,700 | 187 | ~4.8M |
| Contender | Liverpool, Atletico Madrid, Dortmund | ~15,000 | 187 | ~2.8M |
| Standard (mid-table) | Villarreal, Freiburg, Fulham | ~6,300 | 187 | ~1.2M |

These figures are consistent with independent estimates: the Howden Injury Index reported EUR 732M in total injury costs across the Big 5 European leagues in the 2023-24 season (∼EUR 178K per injury), while Nieto Torrejon et al. estimated that the top 5 LaLiga clubs collectively spend EUR 65.4M per year on muscle injuries alone [20]. Beyond salary, competitive revenue effects amplify the impact: each league rank is worth approximately EUR 4.9M in Bundesliga TV and prize money, and missing Champions League qualification costs an estimated EUR 27M [21].

To put these figures in concrete terms: 15 muscle injuries per squad per season at 96% non-contact yields approximately 14.4 addressable injuries per season (15 × 96% non-contact = 14.4 injuries × 13 days = 187 injury days). A 10% reduction would prevent 1–2 non-contact muscle injuries per squad per season, saving approximately 19 player days; a 20% reduction would prevent approximately 3 injuries and 37 player days. For an elite club, this translates to salary savings of EUR 481K to EUR 962K per season (**Table 1b**). However, note that these projections are illustrative. To our knowledge, no randomised trial has demonstrated that model-guided load management reduces injuries, and most models’ low positive predictive value (see Section 3.4) limits the practical scope for intervention.

**Table 1b.** Illustrative salary savings from hypothetical reduction in non-contact muscle injuries.

| REDUCTION | INJURIES<br>PREVENTED | DAYS SAVED | ELITE (EUR) | CONTENDER<br>(EUR) | STANDARD<br>(EUR) |
| --- | --- | --- | --- | --- | --- |
| 5% | ~0.7 | 9.4 | 241K | 141K | 59K |
| 10% | ~1.4 | 18.7 | 481K | 281K | 118K |
| 20% | ~2.9 | 37.4 | 962K | 561K | 236K |

### 3.2 Longitudinal trends in inflammation, immune age, and training load

Before developing the predictive model, we conducted an exploratory analysis of the longitudinal relationships between systemic inflammation, immune age dynamics, training load, and injury across the 26-player cohort during the 2025 calendar year (January–December 2025). A total of 112 peripheral blood samples (median 5 per player, range 2–6) were collected across 8 sampling dates spanning the 2024/25 competitive season, preseason, and 2025/26 competitive season. GPS-derived training load was available for 13,247 sessions (median 636 per player).

**Figure 2A** shows the team-level trajectories of inflammation (inflammation enrichment score ESn, left axis) and external load (Player Load, right axis) across 2025. The mean team inflammation score remained stable across the year (mean ESn 73.7, SD 2.9), while Player Load showed the expected periodisation pattern: sustained levels during the competitive season, a brief decline during the June transition, and a sharp ramp during July preseason. Individual variation was substantial: 11 of 26 players (42%) had at least one high-inflammation sample (above the 90th percentile, ESn ≥ 77.1), with a subset showing recurrently elevated baselines (**Figure 2D**).

**Figure 2.**
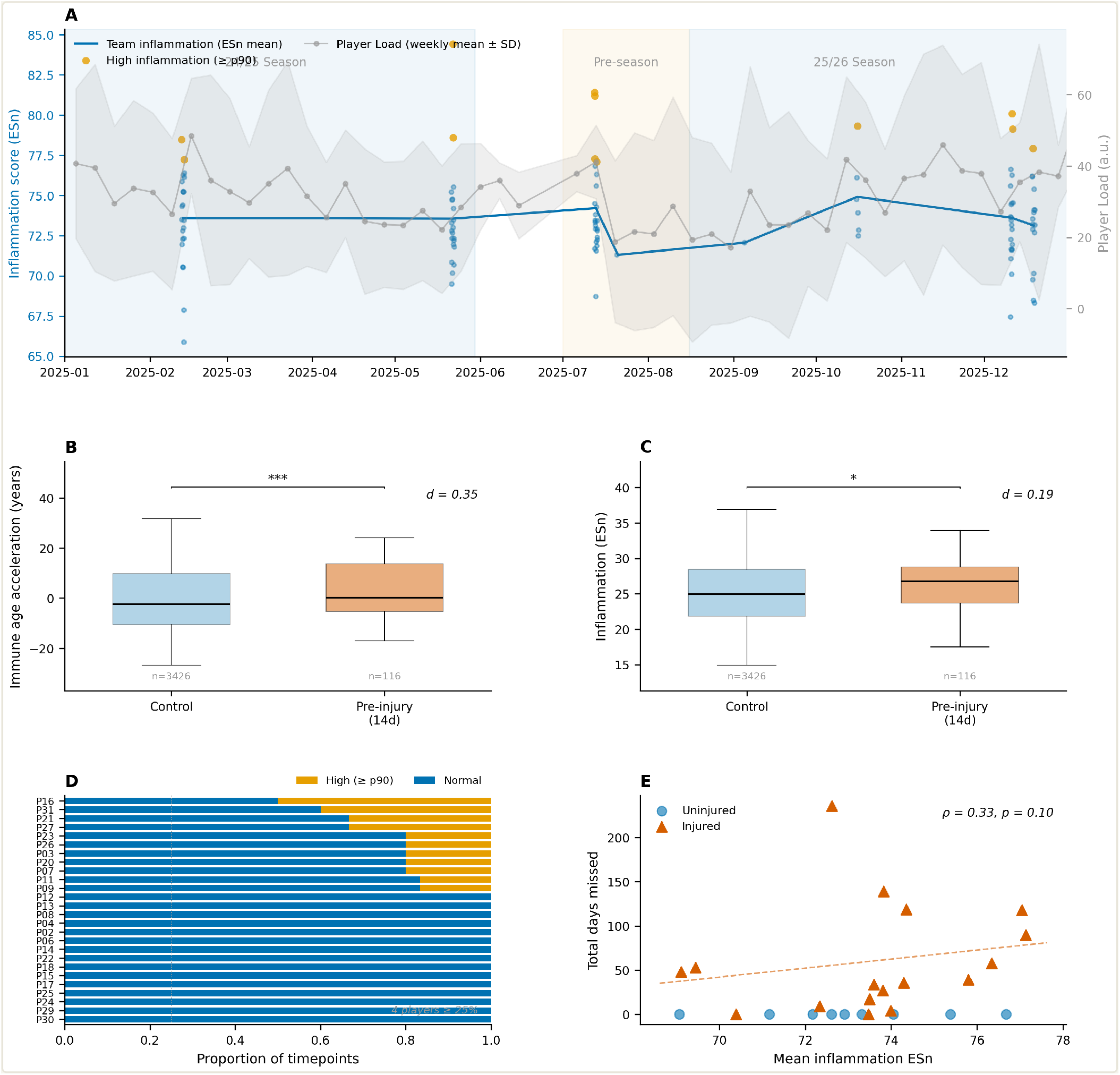
Longitudinal trends and pre-injury feature distributions. (A) Team-level external load (Player Load, grey band showing weekly mean ± SD) and systemic inflammation (inflammation enrichment score ESn, blue line) across the 2025 season. Individual blood samples are shown as dots; orange dots indicate high-inflammation events (≥ 90th percentile). Background shading indicates season phases. (B) Immune age acceleration in control versus pre-injury (14-day window) observations (Cohen’s d = 0.35, p < 0.001). (C) Inflammation ESn in control versus pre-injury observations (Cohen’s d = 0.19, p = 0.048). (D) Proportion of high-inflammation timepoints (≥ 90th percentile) per player, sorted by proportion; dotted line marks the 25% threshold. (E) Mean inflammation ESn versus total days missed per player; triangles = injured players, circles = uninjured; dashed line shows the regression trend for the injured subset (Spearman ρ = 0.33, p = 0.10).

To assess whether inflammation and immune age patterns are associated with injury events, we compared observation-level distributions between the 14-day pre-injury window (n = 116 observations from 15 injury events) and control periods (n = 3,426 observations; **Figure 2B–C**). Immune age acceleration was significantly elevated in the pre-injury window (mean +3.78 years, SD 11.31) compared with control periods (mean -0.89 years, SD 13.33; t-test p < 0.001, Cohen’s d = 0.35; **Figure 2B**). Inflammation enrichment scores showed a smaller but consistent elevation (pre-injury mean 26.20, SD 4.65 vs control mean 25.28, SD 4.93; t-test p = 0.048, Cohen’s d = 0.19; **Figure 2C**). Both effects are summarised in **Table 2**. At the player level, mean inflammation showed a positive but non-significant association with total days missed due to injury (Spearman ρ = 0.33, p = 0.10; **Figure 2E**), a trend driven primarily by two players with both high inflammation and prolonged absence.

**Table 2.**
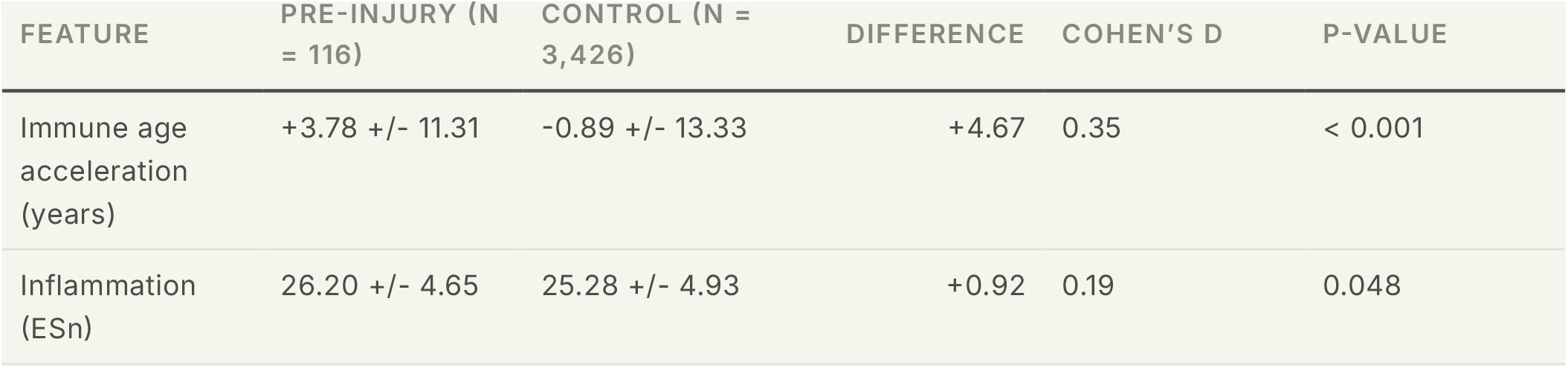
Pre-injury feature distributions (14-day window, observation-level).

The observation-level comparison involves pseudo-replicated daily observations (see Limitations). An event-level analysis collapsing to median values per injury event (n = 15) versus player-season medians (n = 26) showed consistent directions for both features but did not reach significance for inflammation (Mann–Whitney U = 226.5, p = 0.40, d = 0.30), consistent with the low statistical power expected from 15 events.

These exploratory findings: 1) recurrently elevated inflammatory baselines in a subset of players, 2) significantly accelerated immune age in the pre-injury window, and 3) inflammation elevation in this same window; motivated the development of an injury risk model integrating inflammation, immune age, training load, and injury history as features.

### 3.3 Integrating immune monitoring with external load

The injury risk model was built by adding immune monitoring to a GPS-only baseline. Each successive modality improved discrimination, with the fully integrated model raising the overall AUC from 0.541 (GPS only) to 0.678, and the inflammation and immune-age features together accounting for the majority of the gain (**Figure 3A**). Blood biomarkers (creatine kinase, interleuk-in-6, cortisol) were also evaluated but excluded because their sparse temporal coverage reduced the usable dataset from 3,542 to 760 observations.

**Figure 3.**
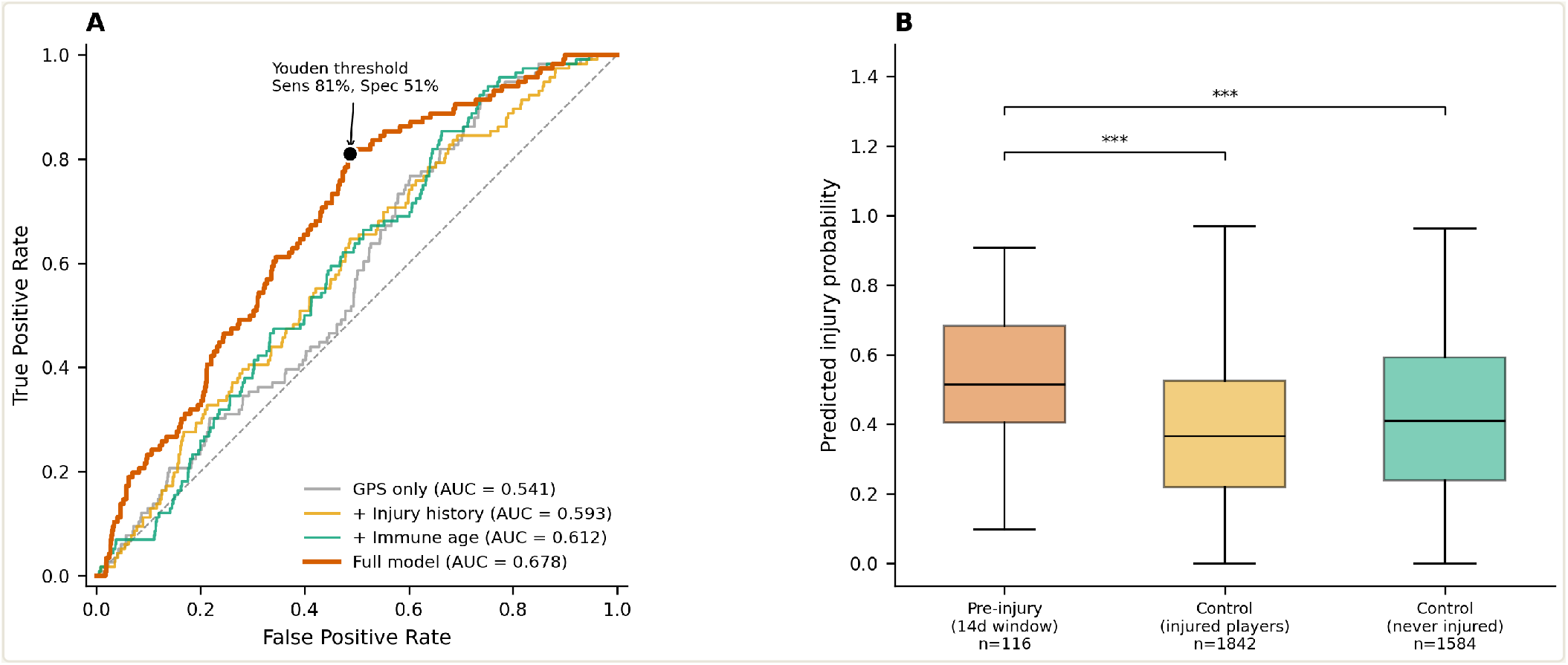
Incremental model performance. (A) Receiver operating characteristic curves for incremental modality addition: GPS-only (AUC 0.541), + injury history (0.593), + immune age (0.612), and the full integrated model adding inflammation (0.678). The black marker shows the Youden-optimal operating point (sensitivity 81%, specificity 51%); the dashed diagonal indicates chance. Each added modality improves discrimination, illustrating that integrating immune monitoring with external load strengthens injury prediction. (B) Predicted injury-probability distributions by group: pre-injury observations (14-day window, n = 116), control observations from the same injured players outside that window, and observations from players who were never injured. Both comparisons against the pre-injury group are significant (Mann–Whitney U, one-sided, p < 0.001).

### 3.4 Model performance

The final integrated model (v2.0, 14-day prediction window) was evaluated on 3,542 daily observations from 26 players, containing 116 positive labels derived from 15 independent injury events (documented via UEFA injury cards). In each of the 26 LOPO folds, the model was trained on 25 players and generated out-of-sample predictions for the held-out player. These per-player prediction vectors were then concatenated across all folds to produce a single set of 3,542 cross-validated predictions, from which the overall ROC curve and all aggregate metrics in Table 3 were computed. Per-player AUC was calculated separately for each fold containing at least one positive observation (AUC range: 0.491–0.947).

**Table 3.**
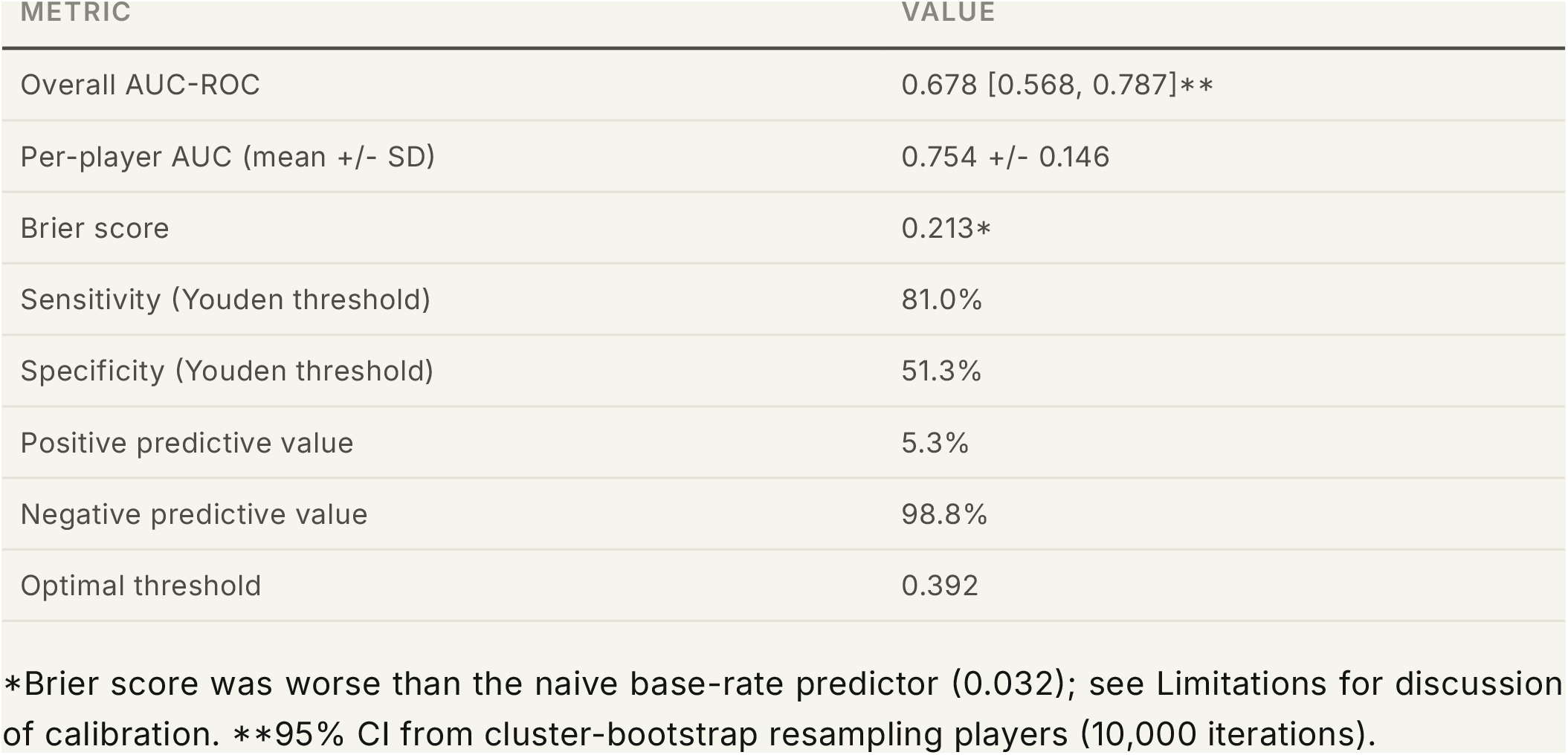
Model v2.0 performance metrics (LOPO cross-validation).

Here, the lower bound of the per-player AUC range (0.491) falls below chance, indicating that the model does not discriminate consistently for all players.

To our knowledge, only two other studies have combined blood biomarkers with GPS-derived load features for injury prediction in football. Rossi et al. [16] reported F1-score 0.63 (injury class) using 7 haematological and hormonal analytes (hematocrit, hemoglobin, RBC, ferritin, sideremia, testosterone, cortisol) in 18 Italian Serie B players, and Haller et al. [17] tested 40 blood biomarkers sampled twice weekly in 25 youth players achieving only 11.1% precision (kappa 0.138), and detecting 1 of 4 test-set injuries. Neither study reported AUCs, precluding direct comparison. With only 4 injury events in the test set against 40 input features, Haller et al. were severely under-powered, making any model’s failure unsurprising regardless of feature type. Whether pathway-level aggregation captures a more stable signal than individual analyte concentrations remains an open question, confounded by differences in population, duration, and sample size between studies (see Discussion). Our GPS-only baseline AUC (0.541) sits at the lower end of the reported literature, but this likely reflects the severity of LOPO-CV without oversampling; the same model class (logistic regression) under random-split validation achieves AUC 0.60 in a comparable study [22].

Predicted risk scores were significantly elevated in the 14-day pre-injury window (median 0.51) compared with both control observations from the same injured players outside the pre-injury window (median 0.37, Mann–Whitney U, p < 0.001) and observations from players who were never injured (median 0.41, p < 0.001; **Figure 3B**). Injured players’ control observations (median 0.37) had lower risk than never-injured players (median 0.41), suggesting that the model assigns reduced baseline risk during non-injury periods for players who were recently injured, potentially reflecting lower GPS load following return from injury.

At a high level, the model’s behaviour reflects two complementary signals: external-load features (notably cumulative weekly load) drive day-to-day fluctuations in predicted risk within a player, while the molecular features (immune age acceleration and inflammation) set a between-player baseline that stratifies players operating at normal versus elevated risk. This decomposition is summarised by per-player SHAP (SHapley Additive exPlanations) contributions [28] (**Figure 4**).

**Figure 4.**
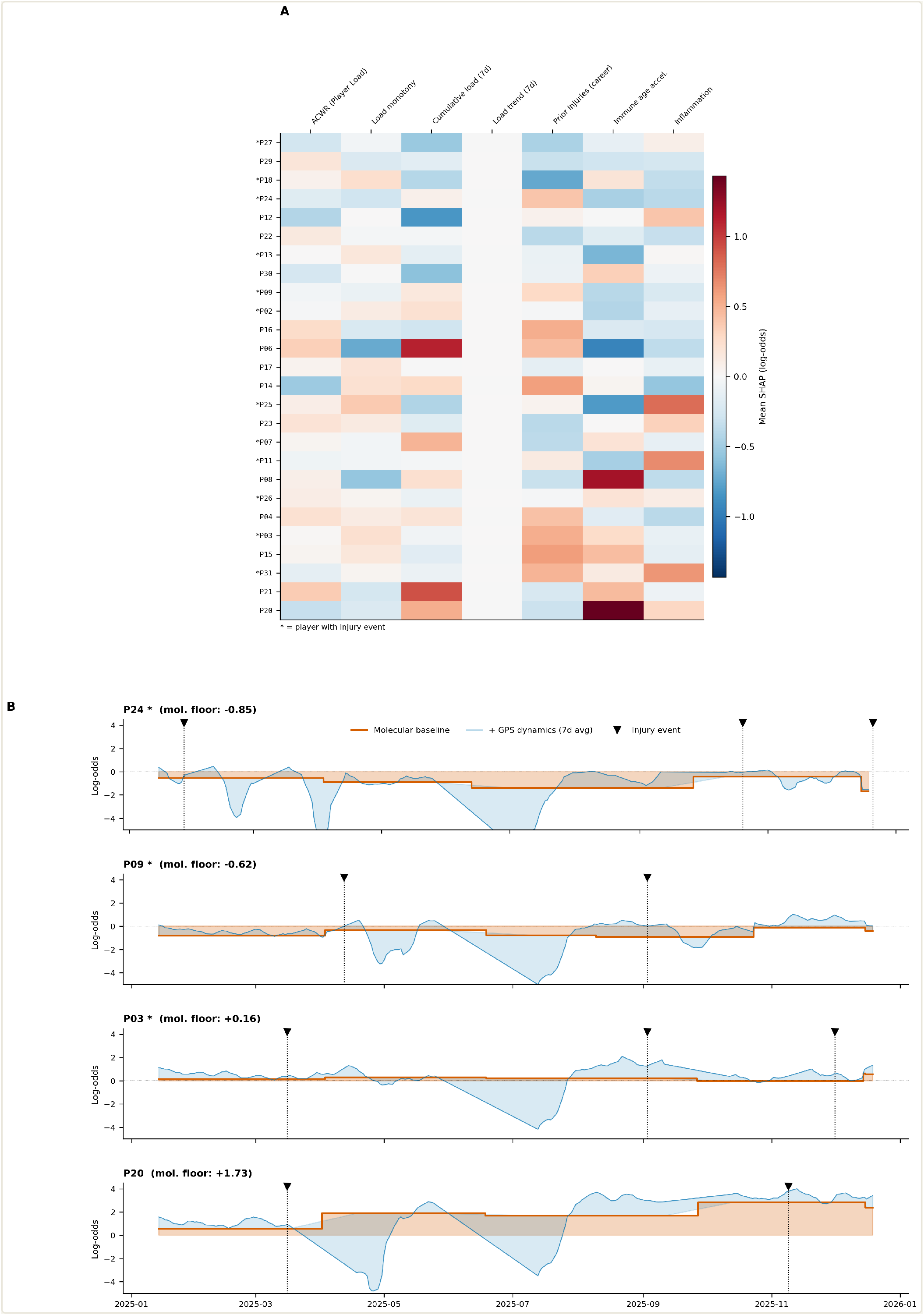
Model interpretation. (A) Per-player mean SHAP heatmap [28]. Rows are the 26 players (ordered by total mean contribution to predicted risk, highest at the bottom); columns are the model’s feature families. The immune-age and inflammation columns show a clear top-to-bottom gradient, indicating that the molecular features drive between-player risk stratification, whereas the external-load features vary mostly within players across sessions. Asterisks mark players with at least one injury event. (B) Temporal risk decomposition for four representative players (ordered by molecular floor). The red step function is the molecular contribution (immune age + inflammation, updating at blood draws); blue shows the 7-day rolling external-load contribution stacked on top. Black triangles mark injury events. The molecular baseline sets a vertical offset between players, while external load drives the day-to-day fluctuations within each player.

The per-player SHAP heatmap (**Figure 4A**) decomposes this further. The immune-age acceleration column shows a clear gradient from low-risk to high-risk players, confirming that immune age drives between-player stratification. By contrast, GPS features (cumulative load, load monotony) vary substantially within individual players across sessions, generating the temporal oscillation visible in risk trajectories (Section 3.5, **Figure 4B**). Several injured players (marked with asterisks) cluster toward the high-risk end, though the pattern is not exclusive, some uninjured players carry comparable molecular risk baselines, consistent with the model’s limited positive predictive value.

The temporal decomposition of individual risk trajectories (**Figure 4B**) illustrates this pattern across four case studies spanning the full molecular floor range. The GPS oscillation (blue) is the noisiest component in every player’s trajectory, consistent with GPS features accounting for approximately 80–95% of within-player temporal variance. However, there is a strong vertical offset between players: P24’s (floor -0.85) entire trajectory lives between -1 and -3 log-odds, while P20’s (floor +1.73) lives between +1 and +3 log-odds. This approximately 2.6 log-odds spread in molecular floor is the reason that immune-age acceleration ranks nearly equal to cumulative 7-day load in mean absolute SHAP impact: both features matter for the prediction, but they contribute through different mechanisms. Tracking a single player over time, GPS features drive approximately 80% of the day-to-day risk signal. Comparing players against each other, molecular features create the stratification between players operating at a normal versus elevated risk baseline, a stratification that GPS load alone cannot explain. P09 (floor -0.62) and P03 (floor +0.16) illustrate intermediate cases: both sustained injuries (black triangles), but P09’s injury occurred only when a GPS-driven spike pushed risk above zero from a low baseline, whereas P03’s moderately elevated molecular floor kept risk closer to the injury threshold even during lower-load periods. P20, despite having the highest molecular floor in the cohort and no injury event in the evaluation period, represents the type of player for whom the model would flag chronic baseline elevation as a risk factor warranting closer monitoring.

### 3.5 Prospective validation

The model produces two complementary views for sports medicine staff. A squad-level risk heatmap (**Figure 5A**) displays predicted injury probability by player and calendar week across the 2025 season. The 15 UEFA injury card events used as model training labels are shown as white diamond markers; additional Transfermarkt-sourced injury events (deduplicated against internal records at a 14-day window) are overlaid as grey triangles to facilitate qualitative visual assessment of risk score patterns around injuries not captured by the internal records. This team-level view highlights periods of elevated squad-wide risk and allows retrospective assessment of whether high-risk cells preceded actual injuries. Individual-level risk trajectories (**Figure 5C**) provide the complementary player-specific view, showing 7-day averaged risk over time for selected case studies.

**Figure 5.**
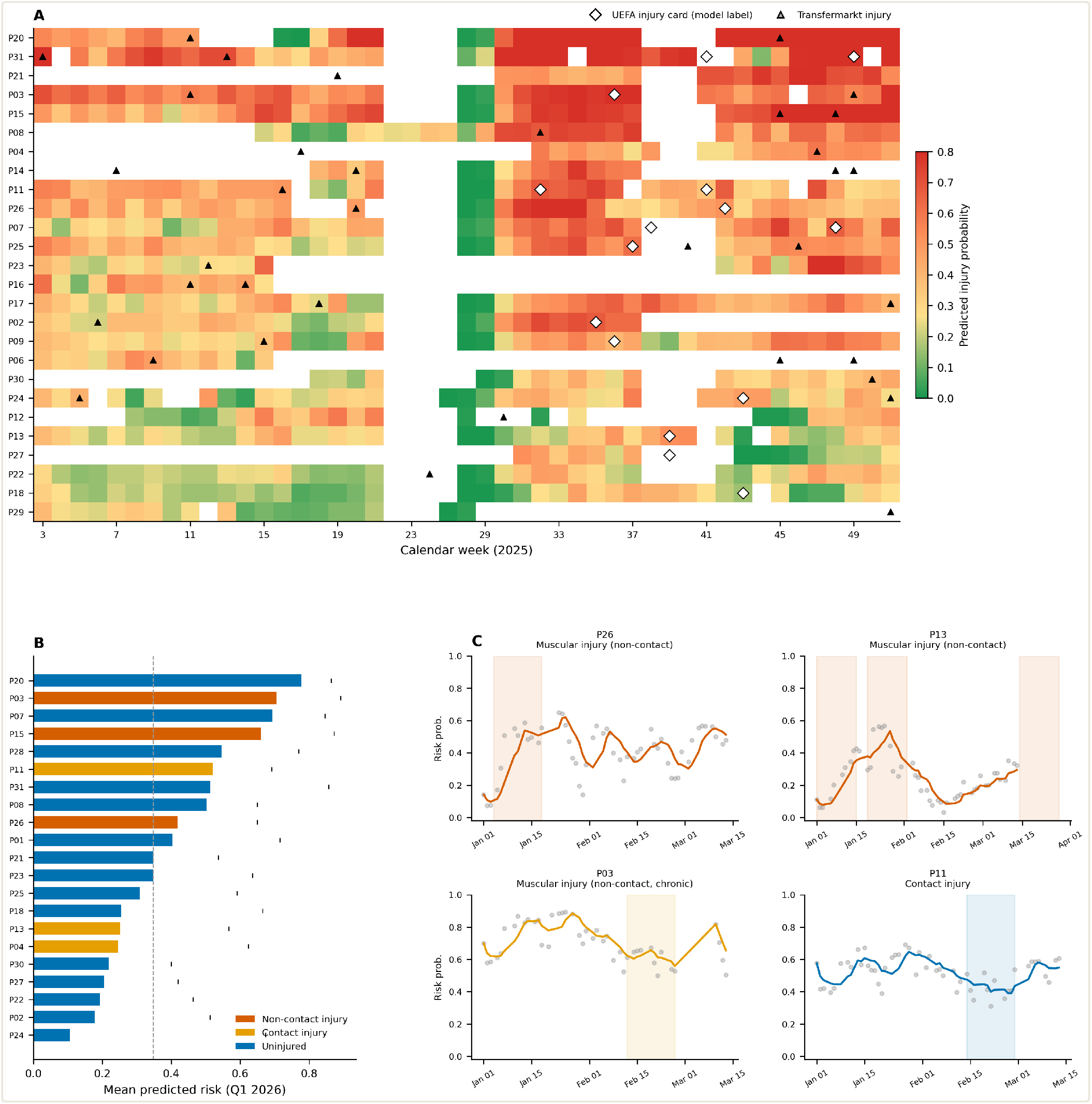
Operational outputs and prospective assessment. (A) Squad-level weekly risk heatmap for the 2025 season. Colour intensity encodes predicted injury probability (0–0.8). White diamond markers indicate the 15 UEFA injury card events used as model training labels; triangle markers indicate additional Transfermarkt-sourced injuries (deduplicated against internal records at a 14-day window) included for qualitative visual context only. Players are ordered by mean predicted risk (highest at top). (B) Q1 2026 donor risk ranking. Mean predicted risk (frozen v2.0 weights) for 21 donors with Q1 2026 GPS data, ranked from highest to lowest. Red = non-contact musculoskeletal injury, orange = contact injury, blue = uninjured. (C) Q1 2026 risk trajectory case studies. Daily predicted risk (grey dots) and 7-day rolling average for four donors. Shaded bands mark the 14-day pre-injury window. Top panels: non-contact muscular injuries with preceding risk escalation; bottom panels: chronic elevation (P03) and contact injury with no pre-injury signal (P11).

To provide preliminary out-of-time evidence, frozen v2.0 model weights were applied to new data from January–March 2026 without retraining. Of the 26 training-set players, 21 had GPS sessions in this window. Feature construction followed the previously described pipeline, with inflammation scores updated to include 8 additional blood samples (sampled in March 2026). In total, 977 of 1,099 GPS-session days had complete feature sets across 21 donors.

Nine injury events were identified across six donors from a public career injury database (Transfermarkt), with an estimated date precision of +/-3–5 days. Events were classified as non-contact musculoskeletal (n = 6) or contact/other (n = 3) based on injury description (**Table 4**).

**Table 4.**
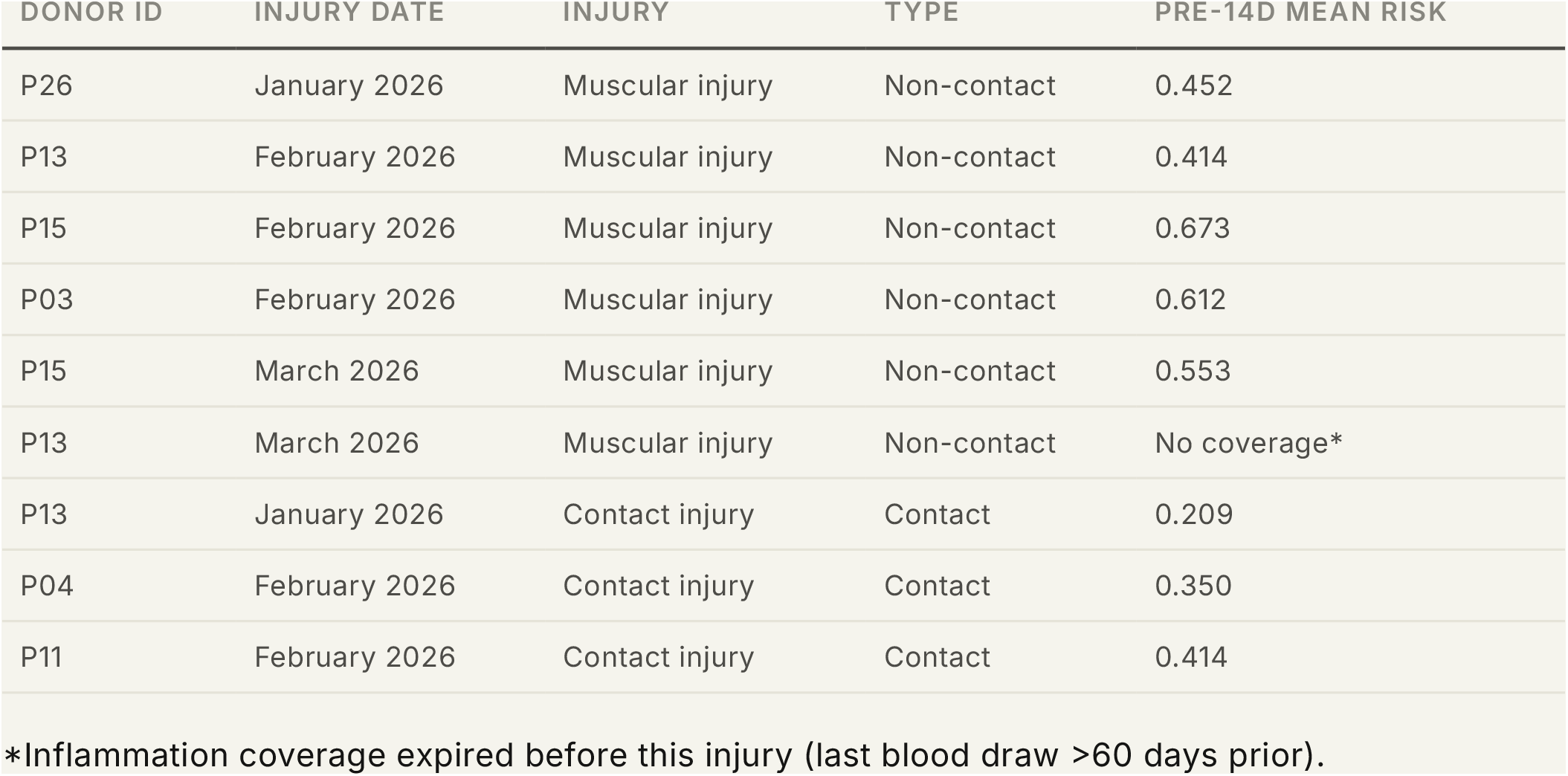
Q1 2026 injury events and pre-injury risk scores.

All injury events in Table 4 are drawn from a public career-injury database (Transfermarkt) and are already in the public domain; the table reproduces no private medical-record information.

At the donor level, players who sustained non-contact musculoskeletal injuries had a higher mean Q1 risk (0.467) than uninjured controls (0.373, n = 15 uninjured donors with predictions; **Figure 5B**). Risk trajectory analysis (**Figure 5C**) revealed two patterns among non-contact injuries: (i) a risk escalation preceding injury, visible for P26 (risk increased from 0.18 to 0.59 over 13 days before his muscular injury) and P13 (risk rose from 0.22 to 0.57 before his February muscular injury); and (ii) recurrently elevated risk throughout the Q1 period, as seen for P03 (sustained range 0.55– 0.67) and P15 (range 0.55–0.79). Contact injuries (P13, P04, P11) were not preceded by elevated model-predicted risk, consistent with the expectation that external trauma is unpredictable from load and inflammation features. P03 exemplifies both the recurrently elevated molecular baseline and the prolonged absence that characterises the high-risk end of the cohort.

Two caveats qualify these findings. First, some uninjured donors had higher mean risk than injured donors (e.g., two uninjured players had Q1 mean risks of 0.694 and 0.777), indicating substantial false-positive exposure. Second, the inflammation feature uses a 60-day matching window, so players whose last blood draw was in mid-December 2025 lost coverage by mid-February 2026. For example: P13’s March muscular injury still fell outside the coverage window of this frozen inference run.

To illustrate the potential practical impact, consider the three Q1 2026 non-contact injuries where the model produced the clearest pre-injury signal. P26’s predicted risk escalated from 0.18 to 0.59 over 13 days before his muscular injury; P13’s risk rose from 0.22 to 0.57 before his February muscular injury; and P03 maintained a sustained risk of 0.55–0.67 throughout the quarter before suffering a muscular injury that resulted in a prolonged absence. In all three cases, model-predicted risk exceeded the Youden-optimal threshold (0.392) well within the 14-day pre-injury window, meaning a load management intervention triggered by the model could, in principle, have been initiated before the injury occurred. Had such interventions prevented even these three injuries, 64 player days would have been saved in a single quarter, roughly equivalent to the epidemiological estimate of 1–2 prevented injuries per squad per full season at a 10% reduction rate (Section 3.1). This is, of course, a best-case counterfactual: we cannot know whether modified training loads would have prevented the injuries, and the model’s low positive predictive value (5.3%) means that many similar alerts during this period would not have preceded an injury. Nevertheless, the temporal alignment between risk escalation and subsequent injury in these cases suggests that the model captures a signal worth further investigation as a complementary monitoring input.

## 4. Discussion

To our knowledge, this is the first study to combine blood-based inflammation profiling and TCR-derived immune age with GPS training load for injury risk modelling in elite football. Integrating the four modalities lifted overall AUC from 0.541 (GPS only) to 0.678, each contributing incrementally, though independence between modalities cannot be confirmed under shared cross-validation folds. Inflammation and immune age acceleration together account for more than half of the predictive performance gain. The underlying biology is plausible: exercise acutely remodels immune and inflammatory signalling in peripheral blood [7,8], with coordinated molecular dynamics persisting over hours to days after a single bout [13]. In a design close to ours, Condeminas et al. [9] linked inflammatory gene expression to GPS-derived external load in competitive handball, showing the load–transcriptome connection in an elite team-sport setting. Peripheral blood inflammation and immune function are therefore exercise-induced and dynamic, with plausible connections with tissue-repair processes relevant to musculoskeletal injury. González et al. [25] extended this to elite female footballers, combining GPS load with multi-omic profiling and identifying tryptophan–serotonin metabolites (serotonin and 5-hydroxytryptophan) as injury-associated. Carmona et al. [26] found that biceps femoris fibres still show focal disruptions 72 hours after a match, indicating that tissue repair often outlasts standard recovery windows. Read together with our results, these studies frame injury vulnerability as an interaction of internal biology and external load.

As a classifier, the model is primarily a rule-out tool (NPV 98.8%), but this should be read against the low base rate: a naive “no injury” classifier already achieves NPV 96.7%, leaving a marginal gain of 2.1 percentage points. Positive predictive value is 5.3% (1,667 false positives against 94 true positives at the Youden-optimal threshold), which limits its value for triggering individual load-management actions. It is better suited to flagging low-risk periods than to triggering interventions.

The Q1 2026 forward inference offers preliminary prospective evidence. Risk rose noticeably before non-contact muscular injuries in P26 and P13, consistent with the model picking up GPS ramps. P03’s recurrently elevated risk suggests that the model also captures long-term vulnerability through career injury count and immune age. This assessment is qualitative: nine events from six donors, with two uninjured donors carrying higher mean risk than most injured players.

These findings are best read as a proof-of-concept for integrating biological internal-load monitoring into elite athlete management. Injury vulnerability emerges from interactions between training load, recovery, immune-inflammatory status, tissue adaptation, and individual biological resilience; immune-inflammatory signatures and immune age acceleration appear to capture part of that signal beyond external load alone. Our work aligns with a multimodal “sportomics” approach: integrating external load, molecular biology, immune profiling, and longitudinal monitoring to characterise the athlete’s internal state and inform individualised care.

Several directions follow. We believe multi-club collaborative cohorts are needed to grow injury counts three-to five-fold and test external validity across competitive levels, sexes, and age groups. Combining immune-inflammatory profiling with other monitoring layers (targeted metabolomics, heart rate variability [15], neuromuscular assessments, sleep, imaging biomarkers, wellness questionnaires, and GPS load) could then help us build individualised biological-readiness profiles, with continuous HRV serving as a daily inflammation proxy between sparse blood draws. A complementary open question is whether persistent immune-inflammatory alterations reflect incomplete tissue recovery, maladaptation to congested fixture schedules, or cumulative biological stress, particularly in recurrent muscle injuries. Here, the longer-term objective would be the development of interpretable monitoring tools that support personalised decisions around recovery, training adaptation, return-to-play, and long-term athlete health.

## 5. Limitations

Several limitations must be considered when interpreting these results.

### Underpowered

The model was trained on 15 internal injury events with 7 features. The events-per-predictor heuristic recommends 70–140 events for stable coefficient estimation with this many features [14]. The reported AUC values and effect sizes are therefore subject to high variance.

### Pseudo-replication of observation-level statistics

The observation-level pre-injury versus control comparison (n = 116 vs 3,426) treats daily measurements as independent units, but these observations originate from only 26 players and 15 injury events; days within a player and within a pre-injury window are not statistically independent. Standard errors and p-values derived at the observation level are therefore optimistically biased. The complementary event-level analysis (Section 3.2, n = 15 events vs n = 26 player-season medians) is provided to mitigate this and shows directionally consistent but non-significant effects, as expected from the low event count.

### Feature selection on evaluation data

Nested cross-validation was infeasible with 15 events; the reported AUC is therefore optimistically biased by an unknown amount.

### Temporal frequency mismatch

The data modalities are sampled at different frequencies (daily for GPS, monthly to bimonthly for blood draws, quarterly for TCR sequencing), creating gaps in individual risk trajectories where molecular features cannot be matched to daily load data.

### Negative ACWR association

The acute:chronic workload ratio was negatively associated with predicted risk, contrary to the conventional U-shaped expectation. We could not resolve whether this reflects survivor bias, fitness confounding, or the model exploiting small within-range fluctuations as a proxy for unmeasured factors; with only 15 events these explanations cannot be distinguished from coefficient instability.

### Calibration

The Brier score of 0.213 is substantially worse than a naive base-rate predictor (Brier = 0.032), indicating poor calibration of absolute risk estimates. With 15 events spread across 26 players, there are insufficient data to fit a reliable probability calibration curve; the model discriminates above chance but does not produce reliable probability scores.

### Single-team, single-sex, elite-only

All data originate from one club’s first men’s team. Training culture, medical protocols, playing style, and the underlying blood signatures themselves may be team-specific. No external validation on other clubs, leagues, or sex was performed. Generalisability to sub-elite, youth, or female athletes is unknown.

## Data Availability

Granular GPS, blood transcriptomic, and immune-repertoire data analysed in this study cannot be shared publicly owing to professional-athlete privacy and contractual obligations. Aggregate results and all performance metrics that support the findings are reported in the manuscript. The proprietary feature-engineering pipeline, molecular feature definitions, and trained model weights are not released in this version and will be described in the peer-reviewed version of this manuscript.

## Declarations

### Ethics approval and consent to participate

The study was approved by the ethics committee of the Barça Innovation Hub (FC Barcelona). All players provided written informed consent in preseason authorising the use of biological, laboratory, and training data for research purposes under the club’s existing medical and performance protocols (see Methods, Section 2.7). No additional procedures beyond routine sports-science monitoring were performed.

### Consent for publication

Not applicable. No individually identifiable data are reported; all players are referred to by anonymised paper identifiers.

### Competing interests

U.P., A.M.-S., M.G., M.S., M.C., and H.H. are affiliated with Omniscope Inc. Omniscope Inc. develops and commercialises the proprietary ImmuneGPT (TCR-derived immune age) and Inflammation Score services that generate two of the model’s input features; ImmuneGPT is the subject of a United States patent application (USPTO Serial No. 99410386). G.R., R.P., L.G.-C., A.M.-G., and M.M. are affiliated with the Medical Department of Futbol Club Barcelona and/or the Barça Innovation Hub.

### Funding

This study was conducted as part of routine sports-science monitoring at FC Barcelona and was supported by the authors’ institutions (Omniscope Inc. and FC Barcelona). No specific external or public grant funding was received.

### Registration and protocol

This was a retrospective observational study; no prospective study protocol was prepared and the study was not registered.

### Patient and public involvement

Patients and the public were not involved in the design, conduct, reporting, or dissemination of this study.

### Data and code availability

See Supplementary Information.

## Supplementary Information

### Code, methods, and data availability

We report the model’s behaviour and performance at a high level and present aggregate results and performance metrics in the manuscript. The feature-engineering pipeline, the molecular feature definitions, and the trained model weights are proprietary to Omniscope and will be described and released in the peer-reviewed version of this manuscript. Granular GPS and transcriptomic data cannot be shared due to player privacy and contractual obligations.

### Proprietary components

The inflammation and immune-age features used by the model are produced by two proprietary Omniscope services, Inflammation Score (a blood inflammation analytics service) and ImmuneGPT (a TCR-derived immune-age model; USPTO Serial No. 99410386), whose internal methods, gene-set panels, and model weights are not disclosed in this manuscript. The external-load and injury-history features follow standard, published methods.

## Notes

### Author Declarations

The study was approved by the ethics committee of the Barca Innovation Hub (FC Barcelona). All players signed informed consent in pre-season authorising the use of biological, laboratory, and training data for research purposes under the club's existing medical and performance protocols.No additional procedures beyond routine sports-science monitoring were performed for this study

